# Cost-effectiveness of polygenic risk profiling for primary open-angle glaucoma in the United Kingdom and Australia

**DOI:** 10.1101/2021.02.18.21251906

**Authors:** Qinqin Liu, John Davis, Xikun Han, David A Mackey, Stuart MacGregor, Jamie E Craig, Lei Si, Alex W Hewitt

## Abstract

**Objective:** Primary open-angle glaucoma (POAG) is the most common subtype of glaucoma worldwide. Early diagnosis and intervention is proven to slow disease progression and reduce disease burden. Currently, population-based screening for POAG is not generally recommended due to cost. In this study, we evaluate the cost-effectiveness of polygenic risk profiling as a screening tool for POAG.

**Methods and Analysis:** We used a Markov cohort model to evaluate the cost-effectiveness of implementing polygenic risk profiling as a new POAG-screening approach in the UK and Australia. Six health states were included in this model: death, early, mild, moderate, severe, and healthy individuals. The evaluation was conducted from the healthcare payer’s perspective. We used the best available published data to calculate prevalence, transition probabilities, utility and other parameters for each health state and age group. The study followed the CHEERS checklist. Our main outcome measure was the incremental cost-effectiveness ratio (ICER) and secondary outcomes were years of blindness avoided per person and a ‘Blindness ICER’. We did one-way and two-way deterministic and probabilistic sensitivity analyses to reflect the uncertainty around predicting ICERs.

**Results:** Our proposed genetic screening programme for POAG in Australia is predicted to result in ICER of AU$34,252 (95% CI AU$21,324-95,497) and would avoid 1 year of blindness at ICER of AU$13,359 (95% CI: AU$8,143-37,448). In the UK, this screening is predicted to result in ICER of £24,783 (13,373-66,960) and would avoid 1 year of blindness at ICER of £10,095 (95%CI: £5,513-27,656). Findings were robust in all sensitivity analyses. Using the willingness to pay thresholds of $54,808 and £30,000, the proposed screening model is 79.2% likely to be cost-effective in Australia and is 60.2% likely to be cost-effective in the UK, respectively.

**Conclusions:** We describe and model the cost-efficacy of incorporating a polygenic risk score for POAG screening in Australia and the UK. Although the level of willingness to pay for Australian Government is uncertain, and the ICER range for the UK is broad, we showed a clear target strategy for early detection and prevention of advanced POAG in these developed countries.

**Copyright:** the Corresponding Author has the right to grant on behalf of all authors and does grant on behalf of all authors, an exclusive licence (or non exclusive for government employees) on a worldwide basis to the BMJ Publishing Group Ltd to permit this article (if accepted) to be published in BMJ editions and any other BMJPGL products and sublicences such use and exploit all subsidiary rights, as set out in our licence.

## INTRODUCTION

Primary open-angle glaucoma (POAG) is the leading cause of irreversible blindness globally,^1–3^ and its prevalence is estimated to range between 2-3% worldwide.^1,4,5^ With an aging population, the projected number of people with POAG globally is anticipated to increase to approximately 112 million by 2040.^5^

POAG has a major disease and economic burden on both individuals and society. Vision loss from POAG is associated with higher risk of falls, depression and unemployment.^6^ Direct healthcare cost estimates for glaucoma were £135 million^7^ across the United Kingdom (UK) in 2002, and AU$144.2 million^8^ in Australia during 2004. It is also well established that the financial burden of glaucoma increases as disease severity increases.^9^ Currently, it is estimated that approximately 50% of glaucoma cases in the developed world are undiagnosed.^4,10,11^ Therefore, early detection of undiagnosed POAG and prevention of progression would be beneficial.

POAG is an ideal disease for screening and risk profiling given it is insidious, leads to significant morbidity if left untreated, and effective treatment is available to slow the rate of progression once diagnosed.^12^ Economic evaluation of any screening programme is essential for government and decision makers due to limited resources. Prior work has shown that the costs associated with POAG are lower when the disease is in earlier stage^13,14^ and, given that advanced POAG at presentation is a major risk factor for legal blindness,^15^ there are clear advantages to screening for POAG. However, previous economic modeling has shown that population-based clinical screening programmes are not currently cost-effective in most high-income countries such as the UK and the US due to high implementation costs and the disproportionate disease burden in the older population.^16^ One potential solution to this predicament is to identify people at high risk for POAG and then undertake targeted clinical screening and monitoring in this high risk subgroup.^16^

Given that POAG is one of the most heritable human diseases,^17^ genetic profiling presents as a unique means to risk stratify and prioritise clinical screening of individuals. People with a family history of POAG have significantly increased odds (between five to 10 times) for developing POAG. Recently, polygenic risk scores (PRS), which combine the weighted risk across a few thousand single-nucleotide polymorphisms (SNPs), has been shown to be a useful tool for estimating an individual’s genetic predisposition to complex diseases such as POAG.^18^ This novel PRS was found to substantially outperform any previous POAG prediction tools based on common genetic variation, whilst also outperforming genetic prediction tools for any other common disease.^19^ Comparing the highest and lowest deciles for the PRS, a 15-fold increased risk for developing advanced disease, as well as an earlier disease onset by up to a decade was identified.^18^ Using the PRS in combination with age, sex and family history to predict POAG diagnosis dramatically outperformed these parameters without the PRS.^18^ As such, PRS profiling could have great health and economic impact on POAG prevention, early detection and intervention for high-income countries.

In this study we sought to determine the cost-effectiveness of implementing a population-wide genetic screening program using a PRS as an adjunct to prioritise individuals for clinical monitoring for POAG. We modeled such a genetic-risk screening programme for the general population aged between 40 to 80 years in the UK and Australia. We believe that this study will provide valuable information for those governments and policy makers such as the Medical Services Advisory Committee in Australia and National Health Service in the UK.

## METHODS

### Study Design

We developed a Markov cohort model to evaluate cost-effectiveness for POAG screening in Australia and the UK from the healthcare payor’s perspective. Based on the mid 2019 population reports by the Australian Bureau of Statistics (ABS), Australia’s national statistical agency,^20^ and statistics from the Office for National Statistics (ONS), the UK’s largest independent producer of official statistics,^21^ we used 11,782,538 Australian and 33,618,730 UK residents aged 40 and above as our study dataset. Individuals with an existing diagnosis of POAG were removed from the sample, based on estimated prevalence and diagnosis rates in the published literature.^4,22^

The proposed design for the screening program is to screen the age groups of interest using a genetic test to obtain the PRS, followed by comprehensive optometrist review every 2 years for those who are identified at high risk via the genetic test. If POAG is diagnosed in an individual, then they enter the established treatment care pathway. The model was built to allow flexibility in determining the population of interest and the PRS threshold for defining individuals at ‘high risk’ and thereby warranting targeted clinical screening.

### Model overview

TreeAge Pro Healthcare^23^ was used to construct the Markov models (Figure. 1), monitor the transition between discrete health states, and represent the cost-effectiveness of the PRS in our population cohorts. Two identical models were built, one for the UK and the other for Australia, utilising different inputs for the key parameters. There were six health states in our model, including healthy individuals, death and four health states associated with POAG stages: early, mild, moderate, and severe. Our definition for the POAG severity classifications was adapted from a prior report^6^ where: 1) “Early” was definite or probable POAG with no visual impairment but changes to optic nerve and/ or retina; 2) “Mild” was definite POAG with mild visual impairment; 3) “Moderate” was definite POAG with moderate visual impairment; 4) “Severe” was characterised by a visual acuity of 6/60 or worse due to POAG.

**Figure 1:**
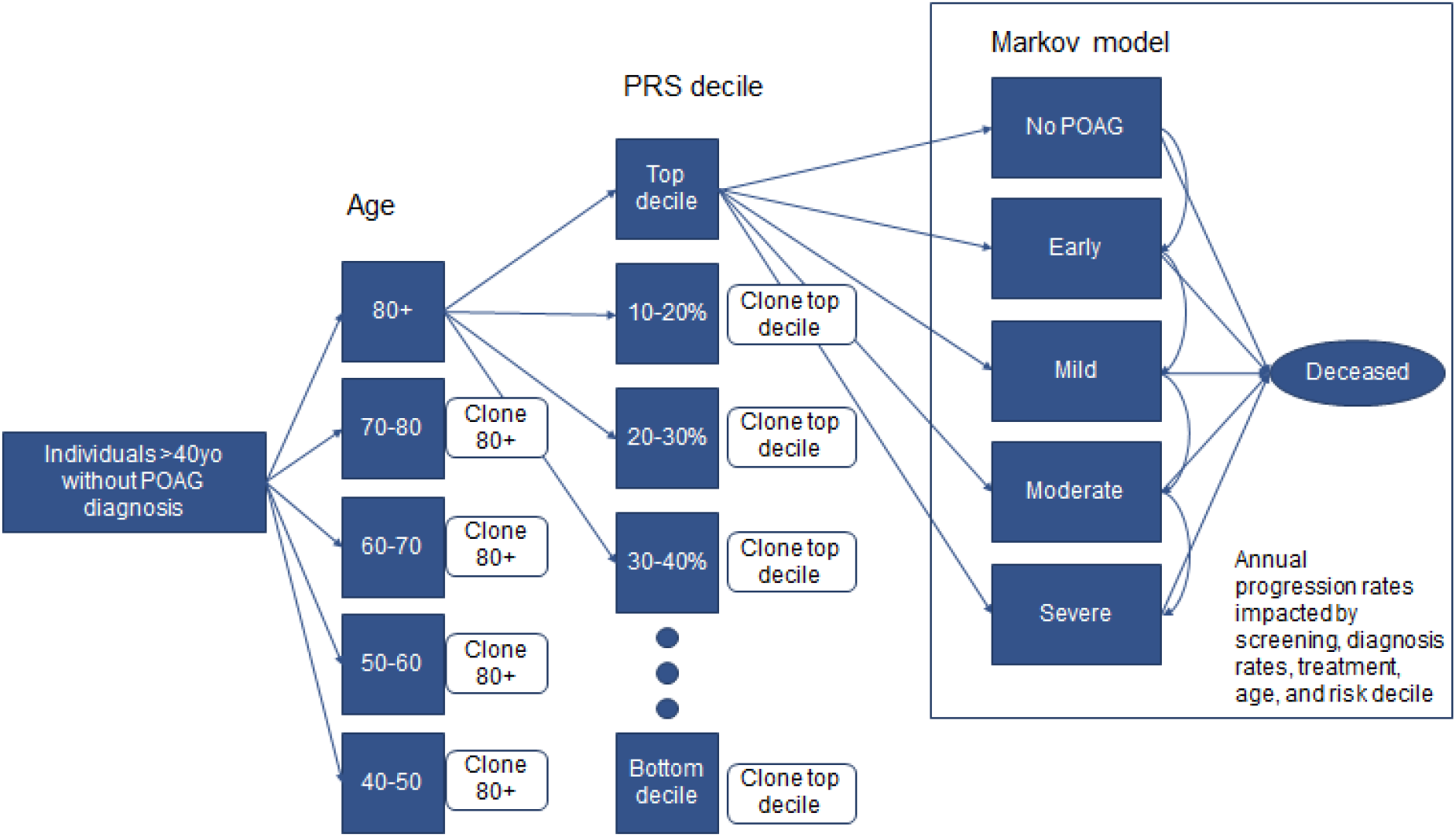
A schematic of Markov Model structure used for the genetic screening of primary open-angle glaucoma ‘pathway’. *i) No POAG: healthy individual; ii) Early: Abnormalities of the optic nerve without any visual field abnormalities; iii) Mild: Damage to the optic nerve and some peripheral vision loss; iv) Moderate: Severe optic nerve damage and advanced peripheral vision loss; v) Severe: Visual acuity of 6/60 or worse or VF constricted to 10°

Our model further divided individuals into age groups (40-49; 50-59; 60-69; 70-79; and 80 and above) as well as into their PRS risk decile. For the base case model, the age groups of 50-59; 60-69 and 70-79 were selected for screening with genetic testing and the top two PRS deciles were also selected for further investigation. A lifetime simulation time horizon was used, whereby individuals could begin the model as either healthy or as undiagnosed POAG at an early, mild, or moderate disease stage based on the prevalence for each disease stage in the Australian and UK population. The methods are stated in the next section. The assumption was made that patients with severe disease would have been diagnosed. Any stage can progress to the next disease stage or to death, but disease stages cannot be skipped nor can a patient remit to an earlier disease stage. The rate of progression to the next disease stage was modified by the individual’s underlying genetic profile (as per their PRS risk decile) and their age and was decreased by treatment.

### Modeling POAG prevalence

After excluding the population with existing diagnosis of POAG, we estimated the prevalence of early, mild, moderate and severe POAG in the screening population within the age groups of 40-49, 50-59, 60-69, 70-79, 80 and above. The following formula was used to calculate the baseline prevalence across a country, age, and disease state level:

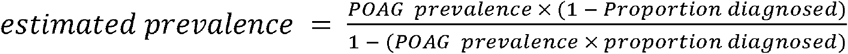

There have been several studies on the prevalence of POAG in Australia and the UK^4,24,22^. After reviewing the available data, the study by Keel *et al*. in 2018^4^, a multi-stage random-cluster sampling with more than 5,000 participants chosen by door-to-door recruitment, was chosen to provide the inputs, based on its methodology, sample size, and recency for Australian data. Keel *et al*. also reviewed whether patients had an existing diagnosis, allowing estimation of diagnosis rates by disease stage. A conservative assumption was made that “probable” POAG in the study by Keel *et al*. aligned with early disease and that “definite POAG” aligned with mild moderate and severe disease. As it is likely that mild POAG was categorised as ‘probable’ in the study by Keel *et al*., for a reasonable proportion of cases, our study underestimates the cost-effectiveness of our interventions.

For the estimation of UK POAG prevalence and diagnosis probability, a meta-analysis by Rudnicka *et al*.^*25*^ from 2006 was utilised to provide the required inputs and adjusted based on the proportion of individuals by race in the most recent data from the UK’s ONS (not shown). On review of the literature, data on disease state as a proportion of POAG diagnosis by age were not available for the UK. Given the cultural, racial, and healthcare system similarities, Australian values were utilised as the base case and then tested in sensitivity testing to understand the ranges for which the results were consistent. Our values for the prevalence of POAG by disease stage by age, in the population without existing POAG diagnosis, are provided in Table 1.

**Table 1.** Summary of the prevalence and transition probabilities for all stages of POAG based on age in Australia and the UK.

| Prevalence at model beginning | Country | 40-49 | 50-59 | 60-69 | 70-79 | 80+ | Source |
| --- | --- | --- | --- | --- | --- | --- | --- |
| No disease | Australia | 99.94% | 98.99% | 98.84% | 97.97% | 97.54% | Calculated from Keel et al.<sup>4</sup> |
| Early | Australia | 0.06% | 0.95% | 0.94% | 1.70% | 1.25% | Calculated from Keel et al.<sup>4</sup> |
| Mild | Australia | 0.00% | 0.06% | 0.03% | 0.04% | 0.15% | Calculated from Keel et al.<sup>4</sup> |
| Moderate | Australia | 0.00% | 0.00% | 0.20% | 0.29% | 1.05% | Calculated from Keel et al.<sup>4</sup> |
| Severe | Australia | 0.00% | 0.00% | 0.00% | 0.00% | 0.00% | Calculated from Keel et al.<sup>4</sup> |
| No disease | UK | 99.71% | 99.46% | 98.99% | 98.04% | 96.24% | Calculated from Rudnicka et al.<sup>25</sup> |
| Early | UK | 0.29% | 0.48% | 0.46% | 0.81% | 1.15% | Calculated from Rudnicka et al.<sup>25</sup> |
| Mild | UK | 0.00% | 0.07% | 0.07% | 0.14% | 0.33% | Calculated from Rudnicka et al.<sup>25</sup> |
| Moderate | UK | 0.00% | 0.00% | 0.49% | 1.01% | 2.28% | Calculated from Rudnicka et al.<sup>25</sup> |
| Severe | UK | 0.00% | 0.00% | 0.00% | 0.00% | 0.00% | Calculated from Rudnicka et al.<sup>25</sup> |
| Annual transition probability | Country | 40-49 | 50-59 | 60-69 | 70-79 | 80+ | Source |
| No disease to Early | Australia | 0.02% | 0.28% | 0.28% | 0.29% | 0.29% | Calculated from Keel et al.<sup>4</sup> |
| Early to Mild | Australia | 0.00% | 3.80% | 9.80% | 10.00% | 17.00% | Calculated from Keel et al.<sup>4</sup> |
| Mild to Moderate | Australia | 85% | 85% | 85% | 85% | 85% | Data from reference<sup>6</sup> |
| Moderate to Severe | Australia | 30% | 30% | 30% | 30% | 30% | Data from reference<sup>6</sup> |
| No disease to Early | UK | 0.03% | 0.14% | 0.14% | 0.22% | 0.90% | Calculated from Rudnicka et al.<sup>25</sup> |
| Early to Mild | UK | 0.00% | 3.50% | 11.40% | 11.40% | 36.50% | Calculated from Rudnicka et al.<sup>25</sup> |
| Mild to Moderate | UK | 85% | 85% | 85% | 85% | 85% | Data from reference<sup>6</sup> |
| Moderate to Severe | UK | 30% | 30% | 30% | 30% | 30% | Data from reference<sup>6</sup> |

### Transition probabilities

All transitions between health states take place in an annual cycle. A thorough search was undertaken on Pubmed and Google scholar to identify transition probabilities by disease stage by age for POAG in Australia and the UK, using the following combined terms: “glaucoma” AND “progression” OR “transition” AND “Australia” OR “United Kingdom”. However, only very limited data were available, and no primary source data were identified at the age/disease stage granularity required for this model. A 50-year annual progression model was created in Microsoft Excel to model population prevalence of ‘early’ and ‘mild and above’ POAG by age group, for a cohort of 40-year-old individuals. Excel’s solver function was used to solve for transition probabilities that met the observed population prevalence (discussed above in ‘Modelling POAG prevalence’), with the constraint that the transition probabilities for each age group must be above zero and higher than the younger age group. Two potential sources were identified as sources for the progression rates from mild to moderate and moderate to severe. The first of these was in the CERA Tunnel Vision study.^6^ The second was “The clinical effectiveness and cost-effectiveness of screening for open angle glaucoma: a systematic review and economic evaluation” by Burr *et al*.,^16^ which utilised visual field mean defect deterioration (MD) and extrapolated it out to more severe disease stages. For this study we utilised the results from the Tunnel Vision study, as it is based on population prevalence, and then investigated these transition probabilities in sensitivity analysis to test model robustness. Transition probabilities are presented in Table 1.

### Utility Values - QALYs and years of blindness

Quality-adjusted-life-years (QALYs) were estimated for population groups from utility data using the following formula:

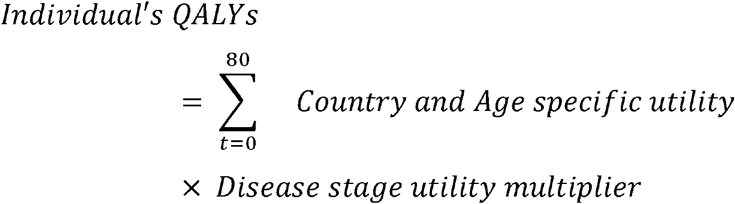

Glaucoma utility multipliers by disease stage were taken from the study by Burr *et al*.^*26*^ for mild and moderate POAG and from Brown *et al*.^*27*^ for severe POAG. No disease and early disease were assumed to have a utility multiplier of 1, and death had a utility value of 0. These disease-based disease multipliers were then applied to utility-by-age data for the countries of interest.^28,29^ The model also evaluated years of blindness, with the disease stage of severe POAG contributing 1 year of blindness per year and all other disease states contributing no years of blindness.

### Screening risk stratification

The recent study by Craig and colleagues^18^ gave the odds ratio (OR) for developing POAG by PRS decile. We adjusted the ORs to be centred around 1 at the population mean risk (Table 2). Then, as the population prevalence for POAG is less than 10%, the rare disease assumption was used to estimate the relative risk (RR) as being the same as the OR. This relative risk was then applied to disease prevalence states at the beginning of the model and to each transition probability to POAG disease states.

**Table 2.** Odds Ratio for each PRS decile in the POAG.

| Decile | Original OR* | Population average-adjusted OR |
| --- | --- | --- |
| 0-10% | 1 | 0.20 |
| 10-20% | 2.33 | 0.46 |
| 20-30% | 1.97 | 0.39 |
| 30-40% | 2.76 | 0.55 |
| 40-50% | 4.04 | 0.80 |
| 50-60% | 3.98 | 0.79 |
| 60-70% | 5.93 | 1.17 |
| 70-80% | 4.87 | 0.96 |
| 80-90% | 8.73 | 1.73 |
| 90%-100% | 14.91 | 2.95 |

\* Original OR: Odds ratio referenced from the study by *Craig et al.*<sup>18</sup>

### Cost

Three distinct cost categories were identified for this model: 1) the cost of the genetic screening test and follow-up monitoring cost for high-risk individuals; 2) the cost of POAG treatment; 3) POAG associated health-economic costs. The details of all cost categories are in Appendix Table 1.

In the base case, screening was assumed at AU$350 and £175 based on the published price of a breast cancer-focused gene panel test in Australia^30^ and the UK,^31^ respectively. The monitoring cost, for those identified as high risk based on their risk decile, was the cost of a follow-up review every 2 years by an optometrist while the individual remained in the ‘no disease’ stage.^32^ All other costs are the cost of POAG treatment by disease stage.

POAG treatment cost for the Australian population was derived from total Australian Glaucoma spending. Glaucoma spending in Australia in 2015, across all categories of spend, was given as $237,208,702 by Australian Institute of Health and Welfare (AIHW).^33^ This was then divided by the estimated number of treated patients based on 2015 ABS age-population data multiplied by the disease prevalence and diagnosis rates published in Keel *et al*.,^4^ to give an estimated $1,867.88 per person-year. Finally, this was adjusted for Australian Health Price Index (HPI) by taking the HPI Compound Annual Growth Rate (CAGR) between 2014 and 2018 and applying it for 4 years to get a value of $2,012.80.^34^ For the UK, the results of work by Rahman *et al*.^*35*^ were utilised and adjusted for UK Consumer Price Inflation (CPI) Index 06: Health^36^ between 2013 and 2020. These values were then adjusted by disease stage based on the ratio of expenditure by disease stage compared to average cost in the study by Varma *et al*.^*37*^

Three major domains were included as POAG-associated health-economic costs based on previous research^38^: aged care facility admissions, falls, and depression. Costs of these events were taken from government data ^33,39^ and other publications^40^ and then applied to the groups based on their age,^39,41^ and glaucoma relative risk.^38^

### Other parameters

#### Diagnosis rates

For the non-screening control model, and for individuals in the screening model who were not in the age groups or risk deciles identified for screening and follow-up, we assumed that the diagnosis rates were the same as those used for prevalence estimation. Patients in the screening model, who were also in the age groups and risk deciles identified for screening and follow-up, had a diagnosis rate of 100%.

#### Treatment impact

Patients who were diagnosed had a reduced risk of progression between disease states (excluding mortality) and incurred full treatment costs. This value was based on Garway Heath *et al*., which demonstrated a Hazard Ratio of 0.44 for patients with glaucoma receiving treatment.^42^

#### Mortality rates

mortality rates were taken by age from the Australian ABS data^43^ and the UK OHS data^44^ and multiplied by relative risk by POAG severity.^6^

#### Willingness to pay (WTP)

WTP for Australia was set at $54,808 based on a parliamentary review into incremental cost-effectiveness ratio (ICER) approved by PBAC for Oncology medication, as no equivalent review was found for ophthalmological treatment or screening programs^45^. In the UK, the WTP threshold is between £20,000 to £30,000, which has been publicly identified by the National Institute for Health and Care Excellence (NICE)^46^ and a value of £30,000 was utilised for this model.

#### Discounting

A global discount rate of 5% p.a. was used for all utility and cost variables in Australia and 3.5% for all utility and cost variables in the UK.

#### Age

All age-dependent variables were based on individuals’ current age in each cycle, not on age at model initiation.

### Outcome

The main outcome was the ICER, defined as incremental cost in the intervention group, divided by incremental QALYs. A positive ICER shows the additional cost per year of QALY gained by the genetic screening program group; a negative ICER demonstrates either a decrease in QALYs or a cost saving (but was not observed in our modelling). A secondary outcome of years of blindness avoided per person, and a ‘Blindness ICER’, defined as incremental cost divided by years of blindness avoided, was also calculated.

### Sensitivity analysis

One-way and two-way deterministic, and probabilistic sensitivity analyses were performed to reflect the uncertainty around predicting ICERs. Variables that had confidence intervals available and were constant between age groups utilised their 95% confidence intervals as their probabilistic parameters. Variables that had confidence intervals available, but are conditional on an individual’s age, used the confidence intervals for the 50-year-old age group, and then multiplied that by the base case ratios between age groups. For cost variables, the study used a range from half to double their values, with the exception of the ongoing monitoring costs for individuals identified as high risk in Australia, which were capped at $150 (representing biannual ophthalmologist review). Transition probabilities that were calculated by the authors or taken from published data without confidence intervals were tested in one-way and two-way sensitivity analyses over arbitrarily large ranges to identify points at which the intervention was no longer cost-effective.

Progression rates from no POAG to early POAG, and from early to mild POAG, utilised beta distributions. The relative risk of POAG based on treatment utilised a lognormal distribution. All costs were modelled on a triangular distribution. ICER uncertainty was evaluated by recalculating the ICER over 10,000 iterations for each country, and the 95% CIs for ICERs were based on the 2.5%-97.5% percentile range of results from the second order Monte Carlo analysis.

## RESULTS

The cost-effectiveness analysis for genetic screening was demonstrated in *TreeAge* as the ICER (Australian dollars and British pounds) and incremental effectiveness (QALYs) (Table 3). The results showed that the genetic screening yielded 1 QALY at a cost of AU$34,252 (95% CI AU$21,324-95,497) and £24,783 (13,373-66,960) per person in our base case in Australia and the UK, respectively.

**Table 3.** Base case cost-effectiveness results from genetic screening for POAG compared to current practice.

|  | Lifetime costs per person, Local currency | QALYs per person | Incremental costs per person, Local currency (95% CI) | Incremental QALYs per person (95% CI) | QALY ICERs, Local currency (95% CI) | Years of blindness per person | Years of blindness avoided per person | Blindness ICERs, Local currency (95% CI) |
| --- | --- | --- | --- | --- | --- | --- | --- | --- |
| **Australia** |  |  |  |  |  |  |  |  |
| Current POAG practice | 53943 | 12.794 |  |  |  | 0.392 |  |  |
| Genetic screening | 54424 | 12.808 | 574 (402-741) | 0.014 (0.006-0.024) | 34252 (21324-95497) | 0.356 | 0.036 (0.015-0.063) | 13359 (8143-37448) |
| **United Kingdom** |  |  |  |  |  |  |  |  |
| Current POAG practice | 36489 | 13.156 |  |  |  | 0.272 |  |  |
| Genetic screening | 37127 | 13.167 | 278 (221-389) | 0.0112 (0.005-0.020) | 24783 (13373-66960) | 0.244 | 0.028 (0.011-0.049) | 10095 (5513-27656) |
| Local currency is Australian dollars for Australia and Great British Pounds for the United Kingdom. Costs, QALYs, and Years of Blindness are discounted lifetime totals per person. ICERs are calculated by comparing the Genetic Screening to the Current POAG practice. QALY=quality adjusted life year, ICER = incremental cost effectiveness ratio, POAG = primary open angle glaucoma. Given values are based on the expected values for the variables, whilst the 95% confidence intervals are derived from the Monte Carlo analysis. |  |  |  |  |  |  |  |  |

Using the WTP thresholds of $54,808 and £30,000, the proposed screening model is 79.2% likely to be cost-effective in Australia and 60.2% likely to be cost-effective in the UK respectively (Fig. 2). The Monte Carlo probabilistic sensitivity analysis showed the screening programme in Australia is likely to be cost-effective in 50%, 80% and 95% of the simulated cases when the WTP threshold is AU$40,608, AU$55,467, and AU$80,170, respectively (Fig. 2a). In the UK model, the screening programme is cost-effective in 50%, 80% and 95% of simulations when the WTP threshold is £27,189, £38,461 and £56,661, respectively (Fig. 2b).

**Figure 2:**
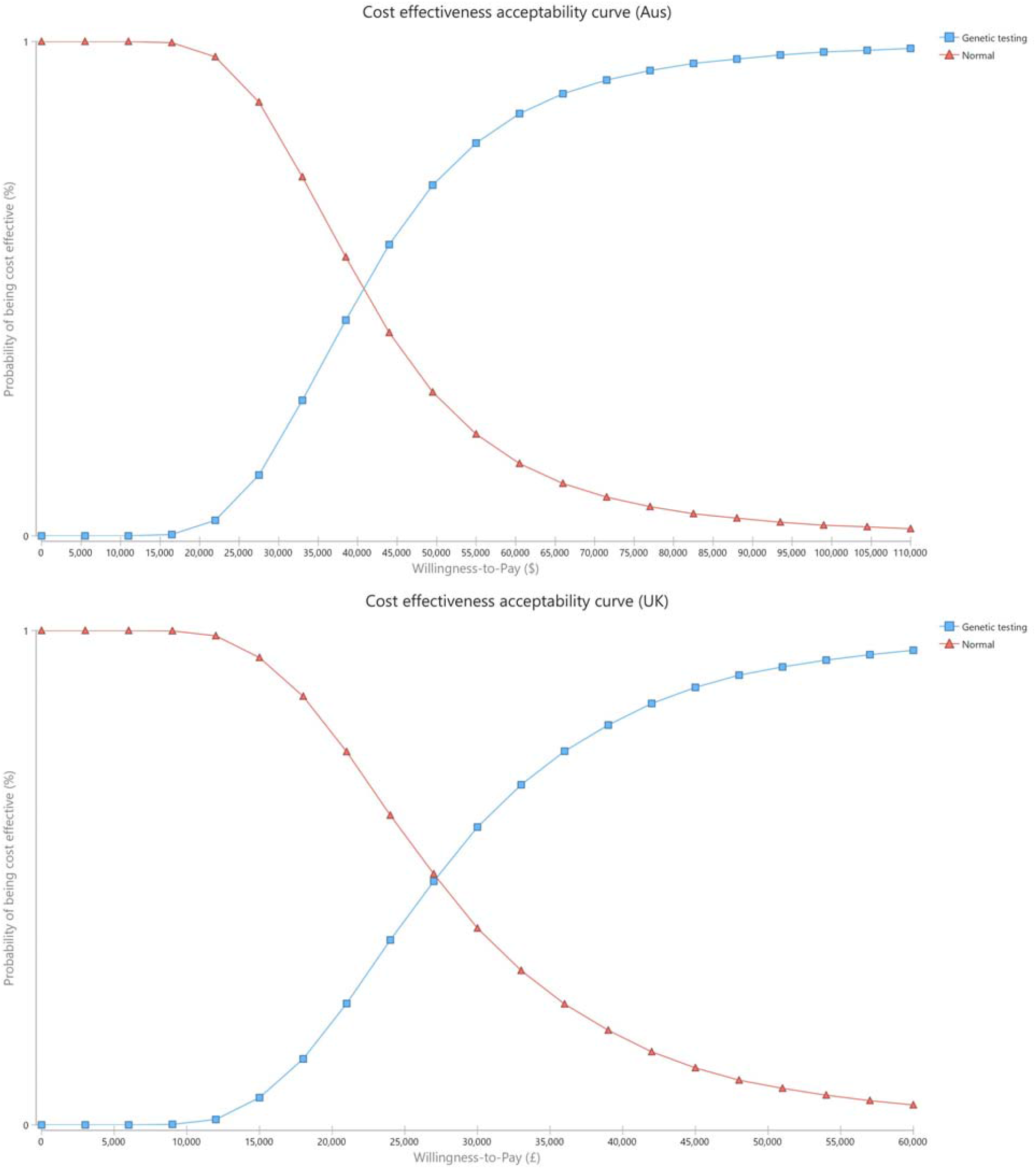
Cost-effectiveness acceptability curve for the genetic screening programme. The probability that the screening programme is cost-effective at different willingness to pay (WTP) thresholds for Australia (a) and the UK (b) models.

In addition, our cost-effectiveness analysis showed that the genetic screening for POAG decreased years of blindness at a reasonable cost. In Table 2, such screening in Australia would avoid 1 year of blindness at ICER of AU$13,359 (95% CI: AU$8,143-37,448), which is one-sixth of the country’s gross domestic product (GDP) per-capita; similarly, it would avoid 1 year of blindness at ICER of £10,095 (95%CI: £5,513-27,656) which is approximately one-third of the UK GDP per capita.

### Sensitivity analysis results

Sensitivity analyses were performed to determine whether the results of the Markov model would change by varying different model parameters. Running the model over 30 years returned an ICER of 34,852 compared to 34,252 for our base model and remained cost-effective (Appendix Table 3). This is as expected, as after 30 years the annual discounting drastically reduces the incremental impact. Furthermore, many age groups have had significant mortality, removing them from the Markov model after 30 years. Similar results were obtained when comparing 30- and 80-year time frames for the UK (not shown).

The tornado diagram displays the parameters with the largest influence on cost-effectiveness (Figure. 3). The top three variables in both countries are the annual screening costs for individuals with high PRS who have not yet developed POAG, the cost of the PRS screening test, and the probability of developing early POAG from no disease.

**Figure 3.**
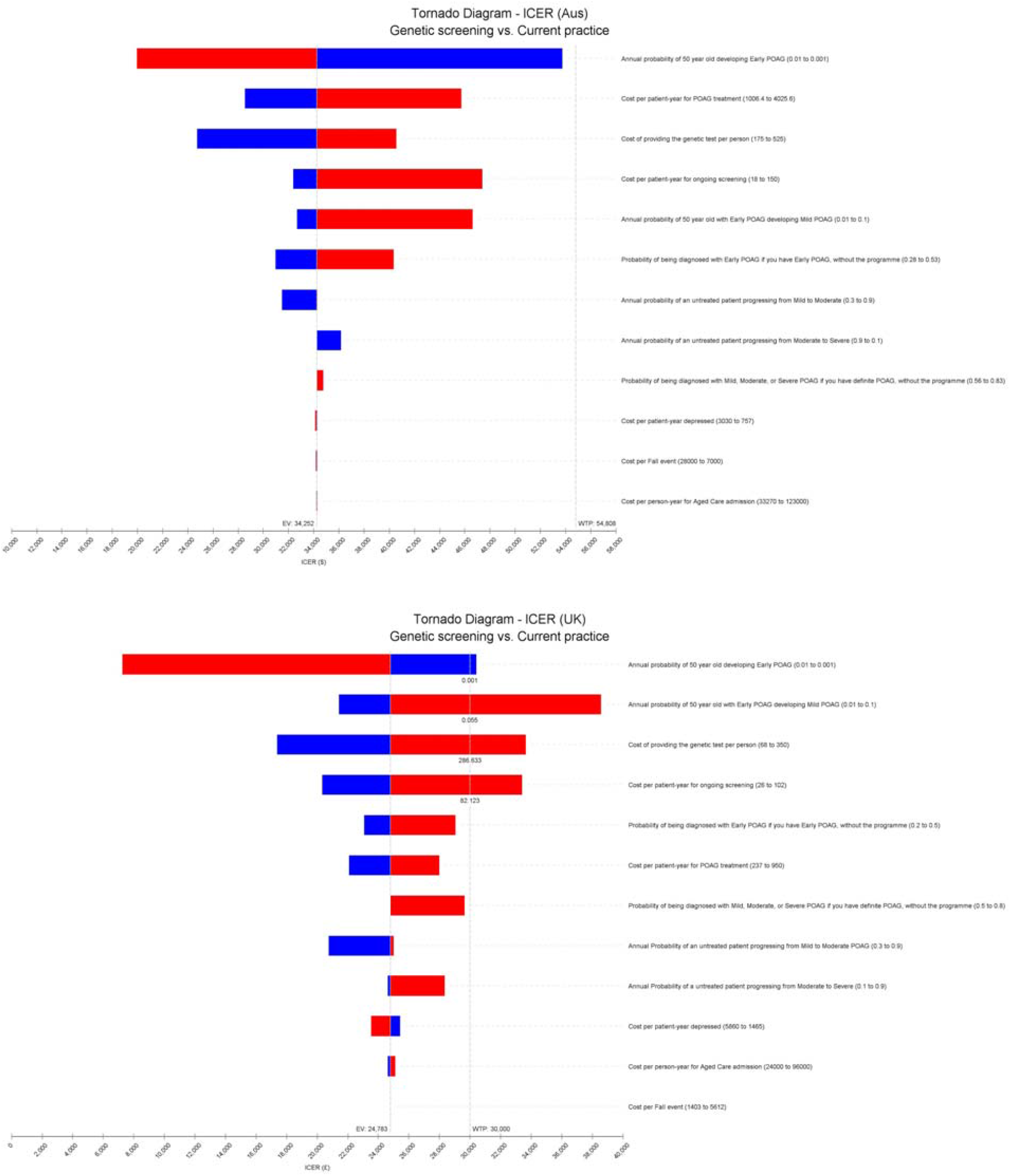
Tornado diagram demonstrating deterministic one-way sensitivity analysis. Costs are given in Australian dollars in Fig 3a and British pounds in Fig 3b. The programme was defined as cost-effective if ICER is less than the WTP threshold, which was set at AU$54,808 in Australia and £30,000 in the UK.

For variables with higher degrees of uncertainty surrounding the data in the literature, we completed sensitivity analyses over arbitrarily large ranges to identify areas where the programme would no longer be cost-effective. With regards to costs, there is a clear pattern between the genetic test and the annual monitoring. In both countries, the cut-off for either cost in terms of generating an ICER below the WTP threshold is co-dependent (Figure. 4a and 4d). It is obvious that the lower both costs are, the more likely the PRS will be cost-effective. We showed that the ICER is below WTP when the incidence of early and transition probability of early to mild are above 0.002 and 0.015 in Australia, and that a similar pattern in the UK model when the incidence of early are above 0.002 and transition probability are above 0.005 (Figure. 4b and 4e). In contrast, our study demonstrated a different pattern between the countries when there is large variation in the transition probabilities of mild to moderate and moderate to severe (Figure. 4c and 4f). In both countries, cost-effectiveness is always below WTP if the transition probabilities of mild to moderate and moderate to severe are both above 0.05; however, in the UK there is an additional constraint where if the transition probabilities are both above 0.6, the screening is no longer cost-effective. Of interest, the region of non-cost effectiveness in the ‘top-right’ of the UK sensitivity analysis for mild to moderate and moderate to severe is cost-effective if the WTP threshold is set arbitrarily higher at £50,000 (not shown).

**Figure 4.**
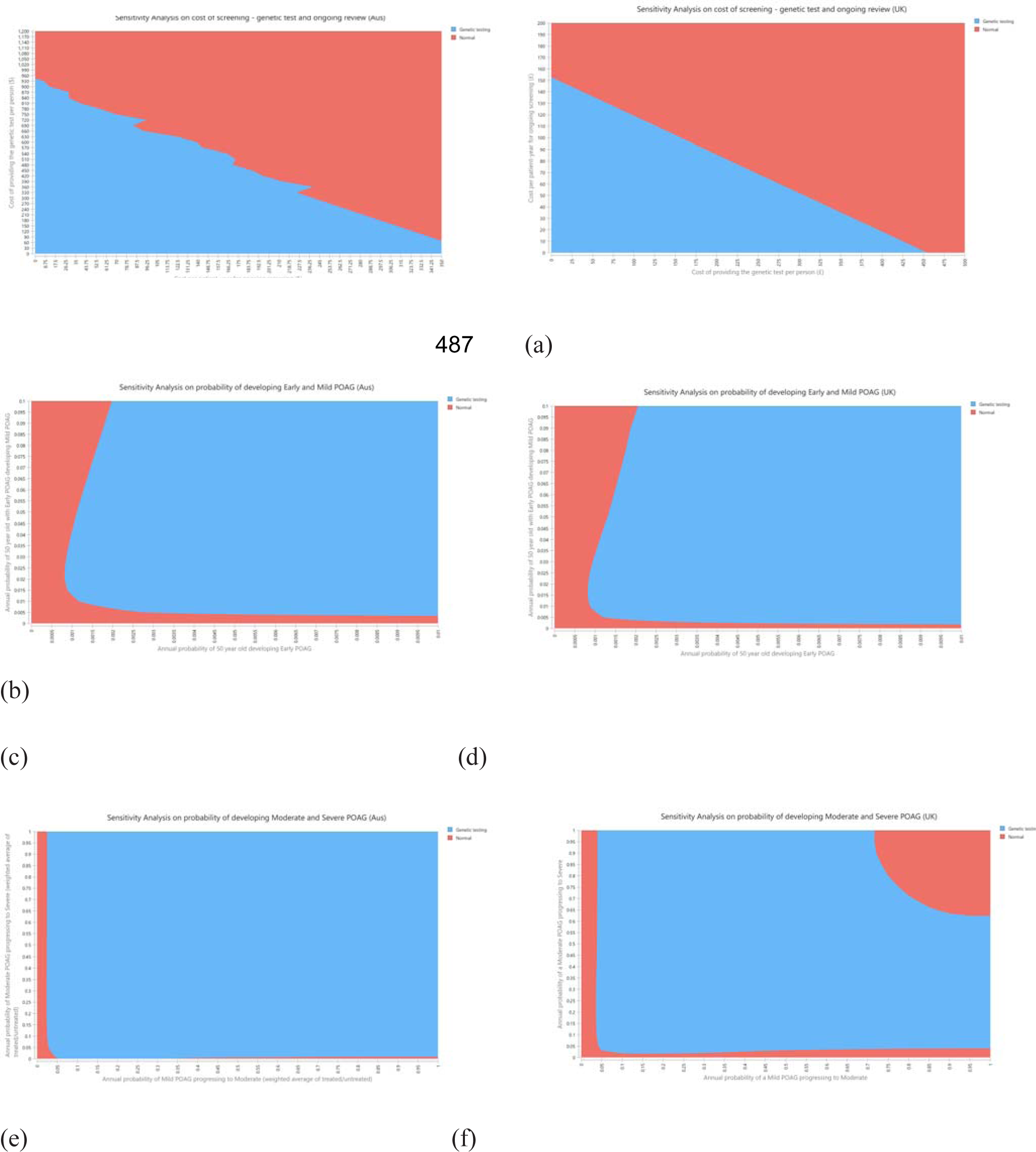
Two-way sensitivity analysis of the cost-effectiveness simultaneously varying key parameters over large ranges. These analyses included cost of screening and cost of genetic test and ongoing review (a, b), probability of developing Early and Mild POAG (c, d), and probability of developing Moderate and Severe POAG (e, f) in Australian and the UK models, respectively.

## DISCUSSION

The World Health Organization (WHO) has established criteria for conditions amenable to screening based on benefit, risk and cost.^12^ According to these criteria, POAG is an ideal target for screening given it is insidious; leads to significant morbidity if left untreated; and effective treatment, by lowering intraocular pressure (IOP) to slow or halt progression, is available.^47^ In 2007, Burr and colleagues published a comprehensive report on cost-effectiveness of screening for POAG in the UK and they concluded that the population-based screening programme for POAG is not cost-effective using strategies such as glaucoma-trained optometrists or dedicated techniques like IOP measurement and visual field test.^16^ To date, no high-income countries have adopted a population-based screening programme for the early detection of the POAG. Although the literature over the last decade has been inconsistent regarding the cost-effectiveness of glaucoma screening in developed countries, the evidence indicates two critical factors influence the cost-effectiveness of a POAG screening programme: 1) screening cost and 2) identification of the high-risk subpopulation.

Our study demonstrated that using the PRS as a POAG screening tool is likely cost-effective for the current Australian population age above 50 and is potentially cost-effective in the UK. Although in the UK model the cost-effectiveness of the genetic testing is cost-effectiveness at WTP or below in 50% of simulations, our ICERs are less than previous UK-based glaucoma screening models.^48^ There are two possible reasons the PRS screening program is more cost-effective when compared to previously proposed screening programmes. Firstly, instead of repeating the screening tests every 3 or 5 years, the single largest cost in our proposed program is a one-off test for each participant and possibly to assess genetic risk for many other diseases.^49,50,51^ Secondly, the PRS would determine a high-risk subpopulation who require regular follow-up, thereby reducing the population who require ongoing repeat screening costs. This would logically reduce the screening cost when compared with repeated IOP measurements, visual field tests and clinical examinations by trained personnel every few years. As a result, our proposed programme would fit the recommendation of targeting high-risk populations at a frequent interval for the POAG screening.

There is growing evidence the PRS could be incrementally valuable to traditional risk factors and implemented clinically to improve current clinical risk prediction models in many common diseases. The U.S. Food and Drug Administration (FDA) allowed marketing of the first direct-to-consumer tests that provide genetic risk information for certain conditions such as Parkinson’s and Alzheimer’s disease in 2017.^52^ Furthermore, research on PRS among the common diseases has expanded exponentially in recent years. Craig and colleagues,^18^ reported substantial evidence of PRS as a remarkable predictor for the risk of POAG progression to advanced POAG, and demonstrates one potential clinical use of PRS – as an adjunct for disease screening. Taking these together, we believe that POAG is a good candidate disease for the first government program utilising PRS screening.

In most high-income countries, optometrists are well-trained to detect POAG. Most previous proposed screening strategies included optometrists conducting the majority of screening tasks. Glaucoma Australia recommended that all Australians 50 years or older visit an optometrist every 2 years for a comprehensive eye exam.^53^ We believe if PRS is implemented for POAG screening, a targeted subpopulation with high risk of developing POAG could safely undergo the same clinical screening. In addition, the subpopulation of individuals with low risk could reduce or delay their monitoring, reducing the total cost of monitoring POAG.

Despite the uncertainty regarding some of the input variables, our sensitivity analysis demonstrated that our results are robust for Australia, with a higher degree of variability in the UK. As this is a new proposed screening methodology, there are no data on the costs for PRS testing for POAG or the annual monitoring cost for healthy but high-risk individuals that this model would identify. Furthermore, the transition probabilities between health states are also not available at an Age by Disease stage level of granularity. The cost of monitoring healthy high-risk individuals has the most impact on our ICER. We believe that the cost would not exceed the standard comprehensive eye exam by trained optometrists in Australia, e.g. less than AU$100 per person per event.^54^ Similarly for the UK, we believe that costs would not exceed £52 every 2 years.^32^ In addition, our sensitivity analysis also showed our ICER was below WTP threshold even if transition probabilities between health states are significantly lower than the evidence suggests. For instance, the PRS programme would be cost-effective even if only 10% of individuals with mild POAG progress to moderate (Fig. 4) per year. According to published reports, the transition probability between mild and moderate POAG is 20% to 85% per year^6,16^ and moderate to severe is 10% to 30% per year.^6,16^ However we acknowledge that our results, in particular for the UK, have a wide range. This could be minimised by the collection of more granular disease severity by age data from the populations of interest.

One of the limitations for this study is the difficulty in determining the true WTP threshold in Australia. In contrast to the UK, which has a published and defined WTP for NHS programs, there is a much wider range of WTP for policy-makers in Australia. For example, according to one recent report published by the Parliament of Australia, the Pharmaceutical Benefits Advisory Committee’s acceptable ICER is in the range of $45 000-$75 000 for cancer drugs.^45^ Given the PRS would be a novel programme for the Australian and the UK Government, the WTP threshold may not be comparable with other drug therapies and interventions. Although our study showed the PRS test would have an ICER of AU$34,252 and £24,783 for each country, this would be the first genetic testing program to be considered as a massive public health prevention programme. The uncertainty of an appropriate ICER for utilising a PRS to reduce vision loss remains in the political domain.

In conclusion, our study was first to report a potential cost-effective POAG screening test using PRS in Australia and the UK. Although the level of WTP for Australian Government is uncertain, and the ICER range for the UK is broad, we showed a clear target strategy for early detection and prevention of advanced POAG in a developed nation.

## Supporting information

CHEERS checklist

## Data Availability

Please contact the corresponding author for all data

## Acknowledgements

D.A.M., S.M., J.E.C., L.S. and A.W.H. are supported by the Australian National Health and Medical Research Council (NHMRC) Fellowships. X.H. is supported by the University of Queensland Research Training Scholarship and QIMR Berghofer Medical Research Institute PhD Top Up Scholarship. We are grateful for funding from a NHMRC Program grant (1150144), Partnership grant (1132454) and a Centre for Research Excellence grant (1116360).

