## Supplementary Material for "Cost-effectiveness of polygenic risk profiling for primary open-angle glaucoma in the United Kingdom and Australia"

**Appendix/Supplements Materials**

**Table 1. Input variables by country for the Markov cohort model**

|  | **Australia value** | **UK value** | **Sources** |
| --- | --- | --- | --- |
| **Proportion of cases diagnosed without screening** |  |  |  |
| Early | 39% | 30% | ^1^^,^^2^ |
| Mild | 69% | 50% | ^1^^,^^2^ |
| Moderate | 69% | 50% | ^1^^,^^2^ |
| Severe | 0% | 0% | Assumption |
| **Transition probabilities** |  |  |  |
| No disease to early disease stage | ***** | ***** |  |
| Early disease stage -> mild | ***** | ***** |  |
| Mild to moderate | 85% | 85% | ^3^ |
| Moderate to severe | 30% | 30% | ^3^ |
| **Treatment relative risk** | 0.44 | 0.44 | ^4^ |
| **Utility multipliers** |  |  |  |
| No disease | 1 | 1 | Assumption |
| Early disease stage | 1 | 1 | Assumption |
| Mild | 0.79 | 0.79 | ^5^ |
| Moderate | 0.64 | 0.64 | ^5^ |
| Severe | 0.26 | 0.26 | ^6^ |
| **Costs (AUD and GBP respectively)** |  |  |  |
| Genetic screening test per patient | 350.00 | 175.00 | ^7,8^ |
| Annual screening costs per patient | 34.43 | 51.28 | Calculated from references^9^ ^10^; |
| Annual POAG treatment costs per patient | 2,012.80 | 563.83 | Calculated from references^11^** |
| Annual depression treatment per patient | 1,405.89 | 2,930.41 | Calculated from references^12,13^ |
| Annual residential aged care facility costs per resident | 66,540.52 | 47,850.90 | ^14^^,^ ^15^ |
| Fall costs per event | 13,466.04 | 2,806.00 | Calculated from references^16,^^17^ |
| **POAG cost weighting by disease stage** |  |  |  |
| Early disease stage | 0.934 | 0.934 | ^18^ |
| Mild | 1.116 | 1.116 | ^18^ |
| Moderate | 1.21 | 1.21 | ^18^ |
| Severe | 1.559 | 1.559 | ^18^ |
| **Relative risk of disease states - Mortality** |  |  |  |
| No disease | 1 | 1 | Assumption |
| Early disease stage | 1 | 1 | ^3^ |
| Mild | 1.67 | 1.67 | ^3^ |
| Moderate | 2.34 | 2.34 | ^3^ |
| Severe | 3.01 | 3.01 | ^3^ |
| **Relative risk of disease states - Depression** |  |  |  |
| No disease | 1 | 1 | Assumption |
| Early disease stage | 1 | 1 | ^19^ |
| Mild | 1 | 1 | ^19^ |
| Moderate | 3.5 | 3.5 | ^19^ |
| Severe | 3.5 | 3.5 | ^19^ |
| **Relative risk of disease states - Fall events** |  |  |  |
| No disease | 1 | 1 | Assumption |
| Early disease stage | 1 | 1 | ^19^ |
| Mild | 1.25 | 1.25 | ^19^ |
| Moderate | 1.49 | 1.49 | ^19^ |
| Severe | 1.74 | 1.74 | ^19^ |
| **Relative risk of aged care facility admission** |  |  |  |
| No disease | 1 | 1 | Assumption |
| Early disease stage | 1 | 1 | ^19^ |
| Mild | 1 | 1 | ^19^ |
| Moderate | 1.8 | 1.8 | ^19^ |
| Severe | 1.8 | 1.8 | ^19^ |
| **Mortality rates by age** |  |  |  |
| 40 | 0.11% | 0.12% | ^20^^,^^21^ |
| 41 | 0.12% | 0.13% | ^20^^,^^21^ |
| 42 | 0.13% | 0.14% | ^20^^,^^21^ |
| 43 | 0.14% | 0.15% | ^20^^,^^21^ |
| 44 | 0.15% | 0.16% | ^20^^,^^21^ |
| 45 | 0.16% | 0.17% | ^20^^,^^21^ |
| 46 | 0.17% | 0.19% | ^20^^,^^21^ |
| 47 | 0.19% | 0.20% | ^20^^,^^21^ |
| 48 | 0.20% | 0.22% | ^20^^,^^21^ |
| 49 | 0.22% | 0.23% | ^20^^,^^21^ |
| 50 | 0.23% | 0.25% | ^20^^,^^21^ |
| 51 | 0.25% | 0.27% | ^20^^,^^21^ |
| 52 | 0.28% | 0.30% | ^20^^,^^21^ |
| 53 | 0.30% | 0.32% | ^20^^,^^21^ |
| 54 | 0.33% | 0.35% | ^20^^,^^21^ |
| 55 | 0.35% | 0.38% | ^20^^,^^21^ |
| 56 | 0.38% | 0.42% | ^20^^,^^21^ |
| 57 | 0.41% | 0.46% | ^20^^,^^21^ |
| 58 | 0.45% | 0.51% | ^20^^,^^21^ |
| 59 | 0.49% | 0.56% | ^20^^,^^21^ |
| 60 | 0.53% | 0.61% | ^20^^,^^21^ |
| 61 | 0.58% | 0.67% | ^20^^,^^21^ |
| 62 | 0.63% | 0.74% | ^20^^,^^21^ |
| 63 | 0.67% | 0.80% | ^20^^,^^21^ |
| 64 | 0.73% | 0.88% | ^20^^,^^21^ |
| 65 | 0.79% | 0.95% | ^20^^,^^21^ |
| 66 | 0.86% | 1.04% | ^20^^,^^21^ |
| 67 | 0.95% | 1.14% | ^20^^,^^21^ |
| 68 | 1.04% | 1.25% | ^20^^,^^21^ |
| 69 | 1.15% | 1.37% | ^20^^,^^21^ |
| 70 | 1.27% | 1.51% | ^20^^,^^21^ |
| 71 | 1.41% | 1.67% | ^20^^,^^21^ |
| 72 | 1.56% | 1.86% | ^20^^,^^21^ |
| 73 | 1.74% | 2.06% | ^20^^,^^21^ |
| 74 | 1.93% | 2.30% | ^20^^,^^21^ |
| 75 | 2.15% | 2.57% | ^20^^,^^21^ |
| 76 | 2.39% | 2.87% | ^20^^,^^21^ |
| 77 | 2.69% | 3.21% | ^20^^,^^21^ |
| 78 | 3.03% | 3.59% | ^20^^,^^21^ |
| 79 | 3.41% | 4.02% | ^20^^,^^21^ |
| 80 | 3.86% | 4.50% | ^20^^,^^21^ |
| 81 | 4.37% | 5.06% | ^20^^,^^21^ |
| 82 | 4.96% | 5.70% | ^20^^,^^21^ |
| 83 | 5.61% | 6.44% | ^20^^,^^21^ |
| 84 | 6.40% | 7.30% | ^20^^,^^21^ |
| 85 | 7.28% | 8.26% | ^20^^,^^21^ |
| 86 | 8.26% | 9.35% | ^20^^,^^21^ |
| 87 | 9.39% | 10.55% | ^20^^,^^21^ |
| 88 | 10.63% | 11.87% | ^20^^,^^21^ |
| 89 | 12.02% | 13.31% | ^20^^,^^21^ |
| 90 | 13.52% | 14.88% | ^20^^,^^21^ |
| 91 | 15.12% | 16.57% | ^20^^,^^21^ |
| 92 | 16.80% | 18.39% | ^20^^,^^21^ |
| 93 | 18.55% | 20.30% | ^20^^,^^21^ |
| 94 | 20.35% | 22.30% | ^20^^,^^21^ |
| 95 | 21.50% | 24.38% | ^20^^,^^21^ |
| 96 | 23.21% | 26.59% | ^20^^,^^21^ |
| 97 | 24.80% | 29.00% | ^20^^,^^21^ |
| 98 | 27.02% | 31.58% | ^20^^,^^21^ |
| 99 | 30.14% | 34.26% | ^20^^,^^21^ |
| 100 | 33.40% | 37.01% | ^20^^,^^21^ |
| *****See Table 2.  **See Method section subtitled *Cost* | | | |

**Table 2. Input variables by age range for the Markov cohort model**

|  | **Country** | **40-49** | **50-59** | **60-69** | **70-79** | **80+** | **Source** |
| --- | --- | --- | --- | --- | --- | --- | --- |
| **Initial state at model beginning** |  |  |  |  |  |  |  |
| No disease | Australia | 99.94% | 98.99% | 98.84% | 97.97% | 0.98% | Calculated from reference^22^ |
| Early | Australia | 0.06% | 0.95% | 0.94% | 1.70% | 1.25% | Calculated from reference^22^ |
| Mild | Australia | 0.00% | 0.06% | 0.03% | 0.04% | 0.15% | Calculated from reference^22^ |
| Moderate | Australia | 0.00% | 0.00% | 0.20% | 0.29% | 1.05% | Calculated from reference^22^ |
| Severe | Australia | 0.00% | 0.00% | 0.00% | 0.00% | 0.00% | Calculated from reference^22^ |
| No disease | UK | 99.71% | 99.46% | 98.99% | 98.04% | 96.24% | Calculated from reference^23^ |
| Early | UK | 0.29% | 0.48% | 0.46% | 0.81% | 1.15% | Calculated from reference^23^ |
| Mild | UK | 0.00% | 0.07% | 0.07% | 0.14% | 0.33% | Calculated from reference^23^ |
| Moderate | UK | 0.00% | 0.00% | 0.49% | 1.01% | 2.28% | Calculated from reference^23^ |
| Severe | UK | 0.00% | 0.00% | 0.00% | 0.00% | 0.00% | Calculated from reference^23^ |
| **Annual transition rates** |  |  |  |  |  |  |  |
| No disease to Early | Australia | 0.02% | 0.28% | 0.28% | 0.29% | 0.29% | Calculated from reference^22^ |
| Early to Mild | Australia | 0.00% | 3.80% | 9.80% | 10.00% | 17.00% | Calculated from reference^22^ |
| No disease to Early | UK | 0.03% | 0.14% | 0.14% | 0.22% | 0.90% | Calculated from reference^23^ |
| Early to Mild | UK | 0.00% | 3.50% | 11.40% | 11.40% | 36.50% | Calculated from reference^23^ |
| **Associated 'cost states' by age** |  |  |  |  |  |  |  |
| Depression | Australia | 7.10% | 4.20% | 4.20% | 2.80% | 2.80% | ^24^ |
| Falls | Australia | 0.00% | 0.00% | 1.00% | 2.00% | 9.00% | ^25^ |
| Aged Care Facility Admission | Australia | 0.00% | 0.10% | 0.40% | 1.80% | 17.50% | ^26^ |
| Depression | UK | 43.00% | 44.50% | 32.50% | 17.00% | 13.00% | ^27^ |
| Falls | UK | 0.26% | 0.39% | 0.69% | 1.58% | 6.72% | ^28^ |
| Aged Care Facility Admission | UK | 0.00% | 0.00% | 0.32% | 1.73% | 8.26% | ^29^ |
| **Utility by age** |  |  |  |  |  |  |  |
| - | Australia | 0.91 | 0.89 | 0.88 | 0.85 | 0.83 | ^30^ |
| - | UK | 0.88 | 0.83 | 0.79 | 0.76 | 0.73 | ^31^ |
| *Insert notes here as relevant* | | | | | | | |

**Table 3. Comparison of running model for lifetime of participants vs. for 30 years**

|  | ICER at lifetime model length | ICER at 30 years |
| --- | --- | --- |
| Aus ($) | 34252 | 34852 |
| UK (£) | 24783 | 26213 |
